# Brief Report: A Phase 1 Study Evaluating Extracorporeal Blood Purification to Remove Circulating Tumor Cells in Patients with Metastatic Pancreatic Adenocarcinoma

**DOI:** 10.1101/2025.04.29.25326588

**Authors:** Susanna Ulahannan, Satish Kumar, Peyton Smith, Jennifer Rios, Kristi Booker, Kayla Martin, Sean Duguay, Pankaj Singh, Lakhmir Chawla

**Affiliations:** Stephenson Cancer Center, University of Oklahoma, OK, USA; University of Oklahoma Health Sciences Center, OK, USA; ExThera Medical, Martinez, CA; Veteran Affairs Medical Center, San Diego, CA

**Keywords:** Extracorporeal blood filtration, Circulating tumor cells, Metastatic pancreatic cancer

## Abstract

**Introduction:** Pancreatic ductal adenocarcinoma (PDAC) is the eighth most common malignancy and patients have a 5-year survival rate of less than ten percent. Early studies suggest that extracorporeal removal of circulating tumor cells and particles may improve both clinical symptoms and cachexia. This study reports the first controlled trial of CTC removal in patients with Stage 4 PDAC.

**Methods:** Five patients were consented under a US FDA investigational device exemption (IDE) for a single treatment of the ONCObind procedure using the Seraph 100 filter. All patients received a double-lumen catheter and underwent an extracorporeal blood purification treatment with the Seraph 100 for 3 hours. Pain scores, circulating tumor cell levels, and erythrocyte sedimentation rates were measured at baseline and post-treatment. Adverse events were carefully monitored during the procedure. At the end of the procedure, the vascular catheter was removed.

**Results:** All patients tolerated the procedure well and no treatment-emergent adverse events were reported during the ONCObind procedure. Patients demonstrated a decrease in CTC levels from a baseline of 3016 ± 1924 cell/mL compared to post-treatment levels of 1410 ± 1564, p = 0.03. The sedimentation rate decreased from a baseline of 41.8 ± 51.0 to a post-treatment level of 29.2 ± 11.6, p = 0.50. The mean pain score improved from a mean of 3.8 ±1.8 to a post-treatment level of 1.3 ± 1.5, p = 0.04.

**Conclusions:** The ONCObind treatment procedure was feasible and well tolerated in a small cohort of patients with metastatic PDAC. Future studies are warranted.

## INTRODUCTION

Pancreatic cancer ranks as the eighth most common cancer by incidence, yet it is the third leading cause of cancer-related mortality.[1] This disparity is largely attributed to the aggressive biology of the disease and the limited therapeutic options available. Most pancreatic cancer cases are diagnosed at advanced stages, unresectable at the time of diagnosis with liver being the most common site of metastasis, with a 5 year survival of less than 10 percent.[2] Treatment options for patients with advanced pancreatic cancer have been limited, primarily consisting of chemotherapeutic regimens.[3-5]

Patients with pancreatic cancer often experience a wide range of debilitating symptoms, including rapid and significant weight loss, abdominal pain, jaundice, nausea, vomiting, and profound fatigue. Pain, which may be caused by the pancreatic mass itself or by liver metastases, is typically severe and can lead to considerable functional impairment, severely affecting quality of life. Approximately 75% of patients with pancreatic cancer experience pain, and over 90% of those in advanced stages suffer from pain that is often difficult to manage, necessitating high doses of opioids that can result in significant adverse effects.[6]

The notion of treating the sequalae of metastatic cancer with extracorporeal blood purification (EBP) to remove malignant cells, particles, exosomes, and other cancer products to improve symptoms and quality of life has been proposed.[7] Recently, Shishido and colleagues demonstrated that in an ex vivo series of experiments Seraph 100 heparin-functional adsorption media can be utilized in blood purification to effectively remove circulating tumor cells from patients with metastatic pancreatic cancer.[8] A follow-up study from this same research group showed that this same heparin media can remove large micro-vesicles and C1Q complement proteins ex vivo from blood of pancreatic cancer patients.[9] Clinical studies in patients with metastatic cancer have been conducted wherein EBP utilizing the Seraph 100 filter have effectively removed circulating tumor cells from multiple types of cancer including pancreatic cancer.[10]

We conducted this Phase 1 open-label single treatment study to determine if the Seraph 100 could be deployed safely in patients with metastatic pancreatic cancer. We sought to determine if the successful removal of circulating tumor cells (CTCs) in these patients may improve clinical symptoms of pancreatic ductal adenocarcinoma (PDAC) and to determine the kinetics of CTC removal to potentially inform longer term studies.

### Methods

#### Trial Design

Open label single treatment safety study. Study was conducted under an open FDA investigational device exemption (IDE -G230144) and registered on clinicaltrials.gov – NCT06481397. The study was approved by the central IRB Advarra (IRB ID #Pro00079800) All patients signed written informed consent prior to undergoing any study procedures.

### Patients

#### Inclusion Criteria

Patients ≥ 18 years of age with metastatic PDAC who experienced disease progression or not tolerating fluoropyrimidine-, oxaliplatin- and irinotecan-based regimens or prior treatment with gemcitabine and nab-paclitaxel or not candidates for chemotherapy. In addition, patients needed to have an ECOG performance status of 2 or less and circulating tumor cells concentration of at least 5 cells/mL.

#### Exclusion criteria

Patients who were pregnant or breast feeding, patients who cannot tolerate placement of dual lumen vascular access, or had platelet counts <50,000, a history of Heparin induced thrombocytopenia (HIT), hemodynamic instability and inability to tolerate extracorporeal procedure, and renal failure.

### Intervention

Before EBP procedure, all patients underwent a complete clinical evaluation including clinical history, physical examination, relevant blood examination and chest/abdominal computed tomography. Local cancer stage was determined according to the TNM classification. After obtaining written informed consent, all patients had a temporary double-lumen catheter placed in the internal jugular vein. Temporary vascular catheter was placed by interventional radiology with the imaging aide and in accordance with standard of medical practice at Stephenson Cancer Center at the University of Oklahoma Medical Center.

The Seraph 100 device was placed in a hemoperfusions mode of the T2008 hemodialysis machine (Fresenius, Bad Homburg, Germany – Figure 1) and with heparin anticoagulation in accordance with standard of medical practice at University of Oklahoma. The setup on the dialyzer machine was for hemoperfusion. Blood flow rate averaged 350 mL/min with an average treatment duration of 180 minutes. After completion of the procedure, the vascular catheter was removed.

**Figure 1:**
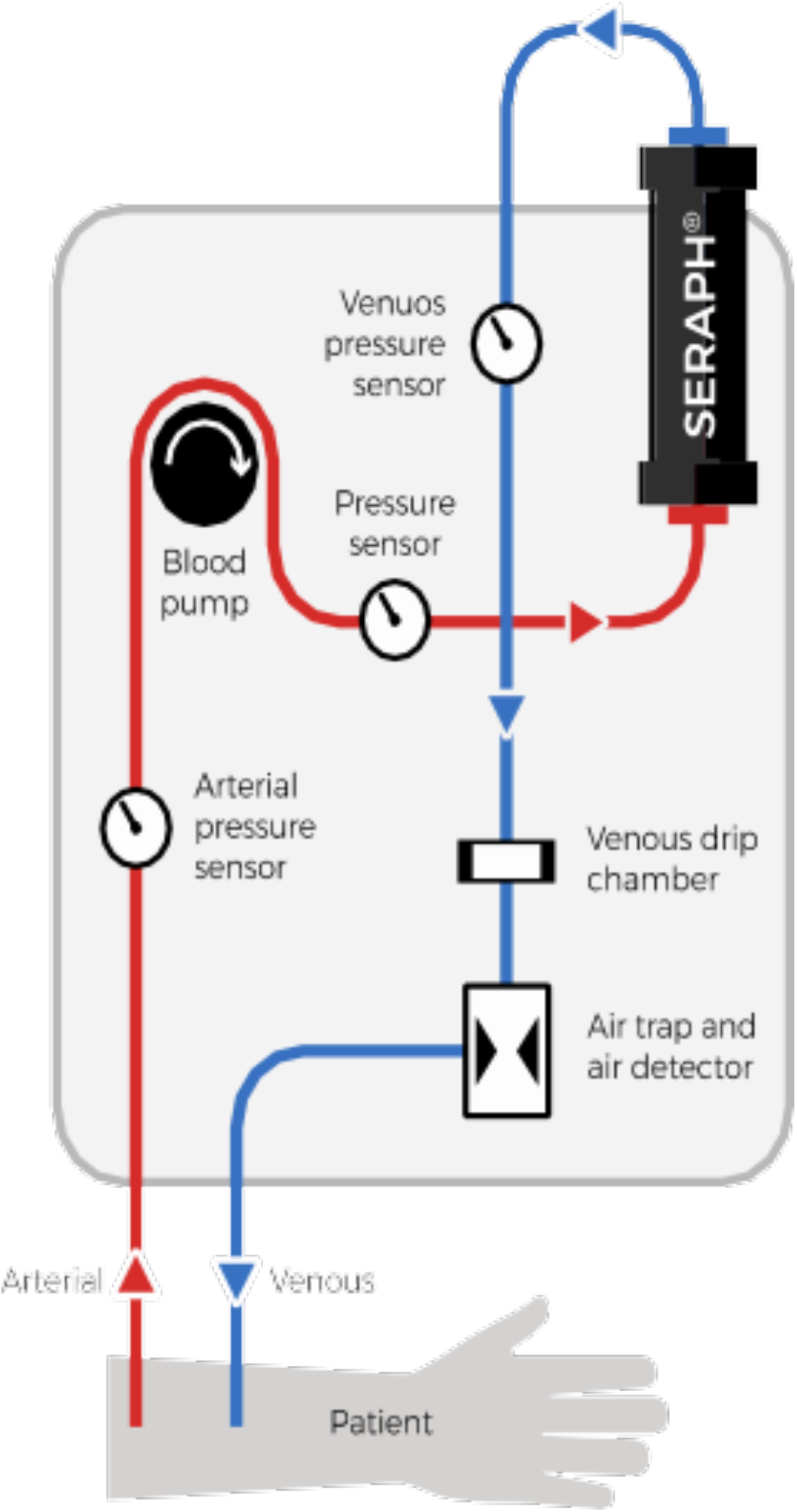
Treatment Schematic

### Assessments and Endpoints

CTC was performed at baseline, 45 minutes, and the end of the session of the EBP session. Both alive cells and dead cells CTC quantification was performed using the Maintrac analysis method (Bayerouth, Germany).[11-13] Determination of CETCs/CTCs in peripheral blood with maintrac® liquid biopsy is based on the maintrac® test system, a microscopy-based detection platform developed by simfo GmbH (Bayreuth, Germany) for the quantification of both live and dead CTCs in patient blood samples.

Erythrocyte Sedimentation Rate (ESR) was measured baseline, 45 minutes after ONCObind procedure commencement as well as at the end of the procedure.

Pain Visual Analog Scale was assessed at baseline, 45 minutes, and at the completion of the EBP treatment.

### Statistical Analysis

Study outcomes are summarized using descriptive statistics. Categorical variables were summarized using frequencies and percentages. Continuous variables were summarized using means, standard deviations, medians, ranges, and numbers of observations.

Baseline CTC concentration levels, ESR levels, and pain score will be compared to post treatment levels with a Sign-E test. For this study, screening and pre-treatment CTC levels were averaged to account for baseline variability. For CTC data that was denoted less than 10, 1 cell/mL was imputed for this reading. For missing data, last value was carried forward for comparative analysis.

## Results

### Patients

Study has enrolled 5 subjects, 3 female and 2 male patients, average age of 60.6 years (min 48, max 76). Four subjects identifying as non-Hispanic White of whom 4 out of 5 were ECOG 1 and 1 patient was ECOG 2. Demographic data is shown on Table 1. All participants provided written informed consent and underwent the intervention between August and September 2024. Baseline targeted physical exams identified common symptoms of metastatic PDAC, including abdominal pain, constipation, diarrhea, and ascites. Vital signs remained stable pre- and post-procedure for all participants

**Table 1:** Patient Demographics.

| Subject | Age Range | Sex | Chemo | Ascites | Level of Metastasis |
| --- | --- | --- | --- | --- | --- |
| 1 | 50-55 | Female | None | Yes | Liver, peritoneum, lung, gastro-hepatic, periportal, retroperitoneal lymph nodes |
| 2 | 75-80 | Female | Gem-Abraxane, ATR Inhibitor and Irinotecan, ADC and Immunotherapy, Liposomal Irinotecan and 5FU | No | Liver, spleen, kidney, lungs, adrenal |
| 3 | 60-65 | Female | FOLFIRINOX, Gem-Abraxane, Polθ Inhibitor and PARP Inhibitor, Liposomal Irinotecan and 5FU, FOLFOX | Yes | Liver, lungs, peritoneum |
| 4 | 60-65 | Male | FOLFIRINOX, Gem-Abraxane, Liposomal Irinotecan and 5FU | Yes | Liver, bone |
| 5 | 45-50 | Male | None | Yes | Liver, omentum and peripancreatic, mesenteric lymph nodes |

### Efficacy Outcomes

The mean live CTC level at baseline, 45 minutes, and post procedure was 61.3 ± 59.8 cell/ml, 33.6 ± 22.5, and 30.4 ± 26.8 cell/ml, respectively. The percentage difference between baseline and post procedure CTC levels was a reduction of 50.4% (p = 0.69), Table 2.

**Table 2:** CTC Levels Before, During, and After ONCObind Procedure.

|  | **Pre-**<br>**Intervention** | **45 minutes** | **Post-**<br>**Intervention** | **Change from**<br>**Pre/Post** | **P value** |
| --- | --- | --- | --- | --- | --- |
| **Live Cells** |  |  |  |  |  |
| Mean (s.d) | 61.3(59.8) | 33.6(22.5) | 30.4(26.8) | 50.4% | 0.69 |
| **Dead Cells** | 3016.7(1924.8) | 1270(1020.2) | 1410 (1564.2) | 53.3% | 0.03 |
| Mean (s.d) |  |  |  |  |  |

s.d. = standard deviation

The mean dead CTC level at baseline, 45 minutes, and post procedure was 3016.7 + 1924.8 cell/ml, 1270 ± 1020.1, and 1410 ± 1564.2 cell/ml, respectively. The percentage difference between baseline and post procedure CTC levels was a reduction of 53.3% (p = 0.03), Table 2.

The mean ESR (sedimentation rate) level at baseline, 45 minutes into the procedure and post procedure was 41.8 ± 51.0, 32.0 ± 11.6, and 29.2 ± 11.6, respectively. The percentage difference between baseline and post procedure ESR levels was a reduction 30.1 % (p = 0.50), Table 3.

**Table 3:** Sedimentation Rate.

|  | **Pre-**<br>**Intervention** | **45 minutes** | **Post-**<br>**Intervention** | **Change from**<br>**Pre/Post** | **P value** |
| --- | --- | --- | --- | --- | --- |
| **ESR** |  |  |  |  |  |
| Mean (s.d) | 41.8(51.0) | 32.0(11.6) | 29.2(11.6) | 30.1% | 0.50 |

s.d. = standard deviation

The mean pain score at level baseline, 45 minutes into the procedure and post procedure was 3.8 ± 1.8, 1.8 ± 2.0, and 1.3 ± 1.5, respectively. The percentage difference between baseline and post procedure CTC levels was a reduction 65.8% (p = 0.04), Table 4.

**Table 4:** Pain Visual Analog Scores.

|  | **Pre-**<br>**Intervention** | **45 minutes** | **Post-**<br>**Intervention** | **Change from**<br>**Pre/Post** | **P value** |
| --- | --- | --- | --- | --- | --- |
| **Pain**<br>**Score** |  |  |  |  |  |
| Mean (s.d) | 3.8(1.8) | 1.8(2.0) | 1.3(1.5) | 65.8% | 0.04 |

s.d. = standard deviation

#### Safety Outcomes

There were no AEs, SAEs, or device malfunctions reported during or immediately after the intervention.

## DISCUSSION

This is the first controlled study of EBP with ONCObind procedure in patients with PDAC. In this Phase 1 study, we demonstrated that EBP to remove CTCs with the Seraph 100 device can be conducted with an acceptable safety profile. Patients showed a reduction in CTCs, reduction in sedimentation rate, and improvement in their pain scores. Additionally, a majority of patients reported an increased appetite, with some patients asking for food immediately after the completion of the procedure.

CTC removal by size exclusion has been attempted previously but has not been successfully deployed into the clinical setting.[14] This size-exclusion technology also utilized an EBP procedure wherein CTCs were removed based on their tendency to be larger in diameter than leukocytes. However, not all CTCs are large, thus limiting the efficacy of this approach. The ONCObind procedure utilizes surface affinity to ‘attract’ CTCs to the filter surface and then trap them on the device surface. Previous studies of the heparin media that is utilized in the ONCObind procedure have demonstrated the capacity to remove CTCs, rare cells, large microvesicles, and complement proteins.[8, 9]

Preclinical studies of CTC removal show potential impact on malignant cells that are not in circulation. Scarberry and colleagues showed the removal of migratory tumor cells impeded metastasis and tumor progression.[15] Similarly, Azarin and colleagues showed that CTC removal via an intra-peritoneal sponge reduced metastasis and a reduction in the primary tumor in the lung.[16] A similar study showed that this same effect could be seen in a pre-clinical study of breast cancer.[17]

In this study, the findings of reduction in CTCs were expected given the previous work that has been published.[8-10] However, the rapid improvement in fatigue, appetite, and pain reduction were unanticipated. These findings were seen across all 5 patients in our study. The precise of mechanism of these observations is unknown. We hypothesize that the removal of cancer cells and exosomes may improve micro-circulatory function thus resulting in the observed findings. In addition, consistent with our observed reduction in ESR, it is possible that the removal of CTCs and other components may reduce inflammation, which is a core competent of cancer-associated cachexia. Future studies of EBP with the ONCObind procedure should assess mechanisms related to pain, cachexia, inflammation, and micro-circulation.

This study had multiple strengths. First, this study was conducted under an FDA IDE with a protocolized treatment regimen. Second, measures of pain, inflammation, and CTC kinetics were performed. Third, the patients were homogenous with metastatic PDAC.

The study also had several limitations. First, the study only assessed a single treatment. Second, measures of inflammation and pain were not measured by more than one type of assessment. Third, measure of microcirculation, functional status, and cachexia were not formally tested. Fourth, the number of patients was modest and the only a single treatment was performed. Lastly, the CTC assay that we utilized has only been validated in research cohorts and is not yet approved assay in the US or Europe.[12, 13]

In conclusion, EBP with the ONCObind procedure in patients with metastatic PDAC appears to be feasible and associated with an acceptable safety profile. Initial observations are encouraging suggesting improvements in pain, appetite, and a reduction in CTC levels. Future studies of the ONCObind procedure in patients with PDAC are warranted.

## References

The authors received no financial support for the research, authorship, and/or publication of this article.

All authors had access to the data and participated in the writing of the manuscript.

## Data availability Statement

All data underlying the results are available as part of the article and no additional source data are required.

Further enquiries can be directed to the corresponding author.

## STATEMENT OF ETHICS Study Approval Statement

Ethical approval was obtained through the Advarra IRB and all patients provided written informed consent

## Conflict of Interest

SU – Advisory Board: Eisai, Astra Zeneca, IgM Biosciences; TAPUR – ASCO Data Safety Monitoring Committee

SK – No COI to report

PS – No COI to report

JR – No COI to report

KB – No COI to report

KM– No COI to report

SD– No COI to report

PS Phd - No COI to report

LSC – Employee of Exthera Medical

Institutional support for research, all funds to institution (Stephenson Cancer Center)

AbbVie, Inc

Adlai Nortye

ArQule, Inc

AstraZeneca

AAtreca

ABoehringer Ingelheim

ABristol-Myers Squibb

ACelgene Corporation

ACiclomed LLC

AErasca

AEvelo Biosciences, Inc.

AExelexis

AExThera Medical

AG1 Therapeutics, Inc

AGlaxoSmithKline GSK

AIGM biosciences

AIncyte

AIsofol

AKlus Pharma, Inc.

Macrogenics

Merck Co. Inc

Mersana Therapeutics

OncoMed Pharmaceuticals, Inc.

Pfizer

Qurgen

Regeneron, Inc.

Revolution Medicines, Inc.

Synermore Biologics Co

Takeda

Tarveda Therapeutics

Tesaro

Tempest

Vigeo Therapeutics Inc.

